# Olfactory, gustatory and trigeminal changes in non-hospitalized COVID-19 patients: an exploratory prospective cohort study

**DOI:** 10.1101/2025.06.20.25329477

**Authors:** Jip M. van Elst, Adriana Tami, Maria F. Vincenti-Gonzalez, Karin Wold, Linda Veloo, Bernardina T.F. van der Gun, Harriët Jager-Wittenaar, Anna K.L. Reyners, Jacco J. de Haan

## Abstract

**Objectives:** This study aimed to longitudinally assess the prevalence and characteristics of olfactory, gustatory, and trigeminal changes, and the impact of these changes on daily life and quality of life (QoL) in non-hospitalized COVID-19 patients.

**Methods:** Three weeks after confirmed COVID-19 diagnosis, non-hospitalized adult participants enrolled in the COVID HOME study consented to complete a questionnaire by phone on the presence, characteristics, and impact on daily life and QoL of olfactory, gustatory and/or trigeminal changes. Participants reporting taste and/or smell changes completed the same questionnaire at three months and, when still present, at six months.

**Results:** The questionnaire was completed by 94/117 participants included in this study three weeks after COVID-19 diagnosis. The 43 participants with smell and/or taste changes completed the questionnaire at three months, and 19 at six months. Of the 94 participants, 56% were female and the median age was 43 [IQR 29-55] years. At three weeks post-infection, olfactory, gustatory, and trigeminal changes were reported by 40 (43%), 37 (39%), and 8 (9%) participants, respectively. During follow-up, olfactory, gustatory, and trigeminal changes were reported by 17 (40%), 14 (33%), and 3 (7%) at three months, and 12 (63%), 9 (47%), and 1 (5%) participants at six months, respectively. Most patients reported an impact of sensory changes on daily life and QoL, mostly describing it as a bit’ or ‘quite a bit’. However, impact did not differ between time points and most participants reported taking no action to cope with these changes. Participants with reported sensory alterations were most interested in professional help at six months.

**Conclusions:** In non-hospitalized COVID-19 patients, the reported prevalence of olfactory, gustatory and trigeminal changes is higher three weeks after infection. Most patients report modest impact on daily life and QoL due to these sensory changes, and a subgroup reported a profound effect.

## Introduction

Olfactory and gustatory changes are present in 30-75% and 20-69%, respectively, of patients with coronavirus disease 2019 (COVID-19) during the acute phase of the disease[1–3]. Furthermore, COVID-19 patients can experience change of chemesthesis (i.e., cooling-, burning-, or tingling-like sensations) and a dry mouth[4,5]. Flavor is the combination of sensory inputs by the olfactory, gustatory and trigeminal system[6]. If one of these components is damaged, the whole experience of flavor, and thereby food enjoyment, is altered.

In patients with COVID-19, olfactory and gustatory changes have been reported four to five days after symptom onset. In most patients, these changes return to normal in the first two weeks[7]. However, olfactory and gustatory changes are part of the post-COVID syndrome, together with fatigue, dyspnea, mental problems and chest pain, among others. Olfactory and gustatory changes have been reported in 31% of COVID-19 patients one year after infection [8,9]. It is yet unknown if all COVID-19 patients will eventually regain normal olfactory and gustatory function. Furthermore, it is unknown which types of mouthfeel and mouth temperature changes are present in COVID-19 patients and, if present, whether these changes are remaining after a longer period of time.

Olfactory and gustatory dysfunction can have a severe impact on daily life and quality of life (QoL)[10,11]. Disability results from cooking difficulties and the avoidance of certain smells. In addition, the reported negative effects on QoL are due to worrying about problems that arise from olfactory dysfunction, for example, the inability to detect rotten food, smoke, gas leaks, or personal hygiene[12]. Strategies to deal with the negative effects mostly concern asking relatives to help detecting rotten food and maintaining personal hygiene[13]. QoL may also be diminished, for example due to the social compound that is part of eating and the joy people experience when eating. Insight in the severity and type of olfactory, gustatory, and trigeminal changes could help COVID-19 patients to anticipate on the mentioned negative effects and give physicians and dietitians more knowledge to help COVID-19 patients with these changes.

Previous studies have mostly focused on hospitalized patients. Importantly, most patients with COVID-19 are non-hospitalized, and infections with the latest SARS-CoV-2 variant, Omicron, following regular vaccinations are associated with less severe symptoms and a lower admission-to-hospital-rate than infections with previous SARS-CoV-2 variants without vaccinations[14,15]. Therefore, it is important to study the course and burden of symptoms in the non-hospitalized COVID-19 patient group.

In this study, we aimed to longitudinally assess the reported prevalence and characteristics of olfactory, gustatory, and trigeminal changes, and the impact of these changes on daily life and QoL in non-hospitalized COVID-19 patients. It was hypothesized that these changes are present in a substantial subgroup of patients, and, when present, in the majority of patients have an impact on daily life and QoL.

## Patients and methods

### Study design and study population

This study was conducted at the University Medical Center Groningen (UMCG) in Groningen, the Netherlands, between October 2020 and November 2021. Participants were recruited between November 2020 and May 2021. The study is a sub-study of the COVID HOME study, which is a prospective cohort study focusing on non-hospitalized COVID-19 patients[16].

Potential participants were identified at the virology facility of the Department of Medical Microbiology and Infection Prevention and at the Municipal Public Health Services (GGD). Individuals who were positively tested with a PCR-test for SARS-CoV-2, and their positively tested household members, were considered eligible for this substudy. Potential participants were contacted by the COVID HOME team, and were explained the study details. Written informed consent was obtained before data collection. For this substudy, potential participants who were 18 years or older, and comprehended Dutch were included. Participants were excluded if they used medication that seriously affect olfactory and gustatory function. Participants could withdraw or unconsent parts of the study. The COVID HOME study was approved by the Medical Ethics Review Board of the UMCG (METc UMCG no. 2020/158, 2020-07-07), complied with the Declaration of Helsinki for Medical Research involving Human Subjects, and was conducted according to the Dutch law (Wet medisch-wetenschappelijk onderzoek met mensen, Algemene Verordering Gegevensbescherming, Wet op geneeskundige behandelingsovereenkomst).

### Data collection

Characteristics of the participants were collected for the main study through questionnaires that included questions regarding gender, age, smoking status, medical history, current medication, and COVID-19 related symptoms (including anosmia and ageusia).

Participants were called by the researcher three weeks after they had a positive RT-PCR diagnosis for SARS-CoV-2, and if they consented to be called for this substudy. They were asked to answer questions on the presence, characteristics, and impact of olfactory, gustatory and trigeminal changes by phone. The answers were filled in by the researcher in REDCap (Research Electronic Data Capture, Version 10.0.23), a safe, web-based application hosted by the UMCG that supports data-capture. Authors had access to information that identified individual participants during data collection.

Completion of the questionnaire took approximately 15 minutes. Participants reporting olfactory and/or gustatory changes were called again at three months and, when changes were still present, for the third time at six months after diagnosis to fill in the same questionnaire. At the end of each phone call, participants with olfactory and/or gustatory changes were asked for consent to be called again. If participants still reported changes at six months, they were offered dietary counselling by a dietitian with expertise in olfactory, gustatory and trigeminal dysfunction. Participants who could not be reached at one of the time points after multiple phone calls were not called again at the next time points.

### Questionnaire

Apart from the above mentioned questionnaire in which patient characteristics were collected at baseline, a questionnaire previously applied in patients with COVID-19 and patients with cancer was also used in the present study to investigate olfactory, gustatory and trigeminal changes (mouthfeel and mouth temperature) and their impact on daily life and QoL (S1 Questionnaire)[17–19].

In this four-part questionnaire, all participants had to answer questions in the first part regarding alterations in smell, taste, mouthfeel (e.g. dry mouth, tingling sensation, texture) and mouth temperature. If alterations were present, the impact of these alterations on daily life and quality of life was asked on a 4-point Likert scale (not at all, a bit, quite a bit and very much). Impact on daily life refers to how something affects a person’s everyday activities and routines, while impact on QoL encompasses a broader assessment of overall well-being and satisfaction with one’s life circumstances. The second and third part contained specific questions on olfactory and gustatory changes, respectively, and were filled in by participants who reported to experience these changes in the first part of the questionnaire. In both parts, the level of severity of smell or taste perception was asked with the 4-point Likert scale followed with a question on the course of the changes with a 5-point Likert scale (decreases, decreases a bit, stable, increases a bit, increases). Both parts also consisted of open questions concerning coping with smell or taste changes. In the second part specifically, questions on nasal congestion and specific odors were asked. In the third part, changes in intensity in the specific basic tastes salt, bitter, sweet, and sour were described with a 5-point Likert scale (much weaker, slightly weaker, no change, slightly stronger, much stronger). If changes in a specific basic taste were present, the impact of and coping with these changes was asked with the previously mentioned 4-point Likert scale and an open question, respectively. At last, questions regarding metallic taste and continuous taste were asked. The fourth part of the questionnaire consisted of one question: if participants would like more guidance for their changes in smell or taste, which was answered by participants with olfactory and/or gustatory changes.

### Statistical analysis

Statistics were descriptive and data are reported as number with percentage or median with interquartile range. The data was analyzed with IBM SPSS Statistics 23 (IBM Corp., Armonk, NY). Pairwise deletion was applied with missing data.

## Results

### Study population

A total of 193 SARS-CoV-2 positive individuals were included in the COVID HOME study. Of the 193 participants, 27 were children, 31 withdrew from the study, 16 did not want to be called for this substudy, and two did not speak Dutch. Therefore, in this substudy, 117 participants were enrolled between October 2020 and May 2021. Of these participants, 17 did not respond and six participants could not be called timely. At three weeks, 94 participants were interviewed and of these, 45 participants that reported olfactory or gustatory changes at three weeks were called again at three months. Next, 19 participants with olfactory and/or gustatory changes at three months were called a last time at six months. Two participants could not be contacted at three months (Fig 1). No participants were excluded. Of the 94 participants, 53 (56%) were female and the median age was 43 years [IQR 29-55]. Characteristics of the study population are summarized in Table 1. All relevant data can be found in S1 Data.

**Fig 1.**
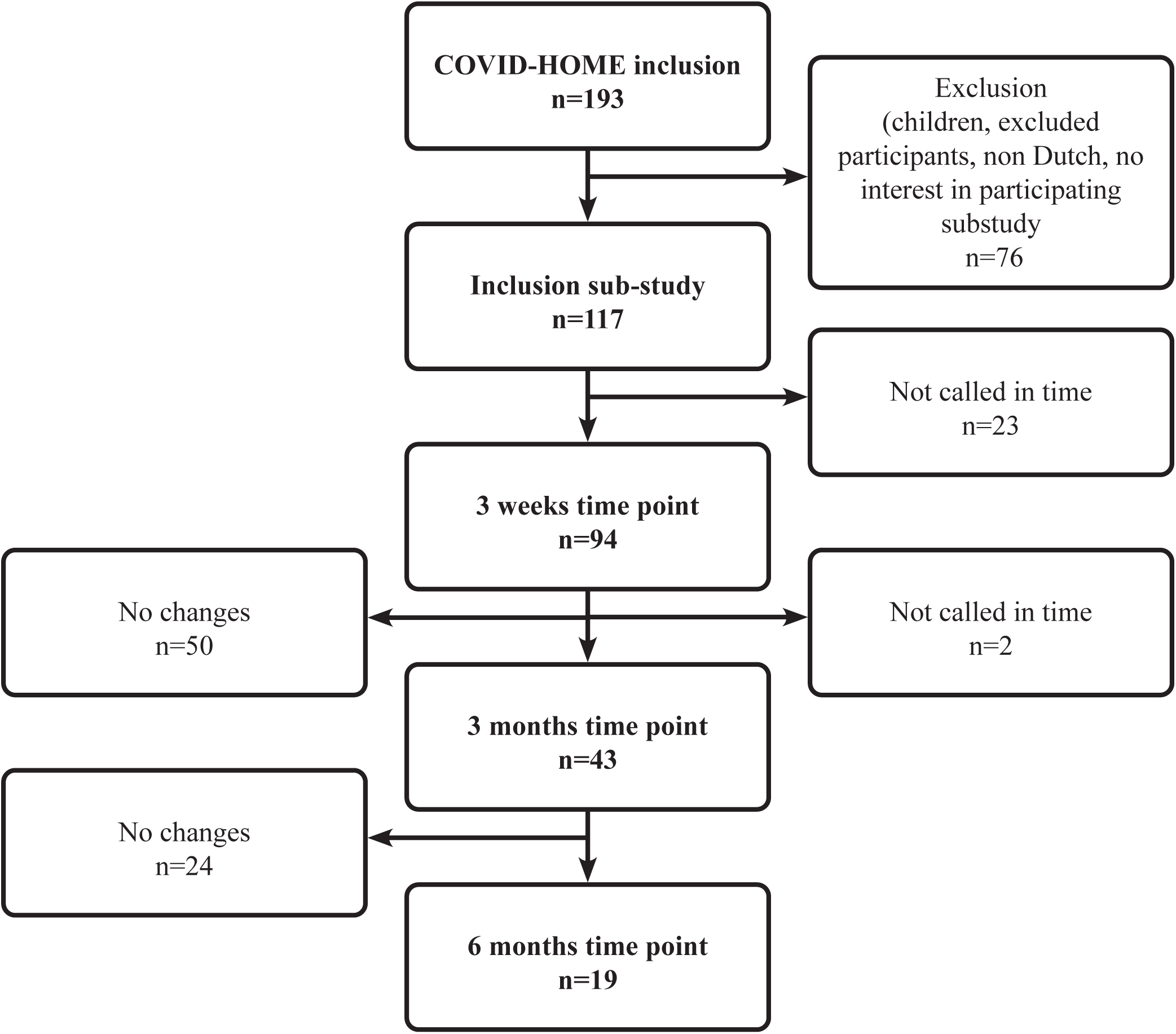
Inclusion of COVID-HOME participants (n) with COVID-19 infection.

**Table 1.** Baseline characteristics.

| Characteristics | Total<br>(n=94) |
| --- | --- |
| Gender, n (%) |  |
| →*Female* | 53 (56.4%) |
| Age (years), median [IQR] | 43 [29-55] |
| Nationality*, n (%) |  |
| →*Dutch* | 85 (95.5%) |
| →*Other* | 4 (4.5%) |
| Smoking*, n (%) |  |
| →*No* | 54 (63.5%) |
| →*Yes* | 7 (8.2%) |
| →*Former smoker* | 25 (29.4%) |
| COVID-19 treatment, n (%) |  |
| →*None* | 75 (79.8%) |
| →*Paracetamol* | 16 (17.0%) |
| →*Antibiotics* | 2 (2.1%) |
| →*Inhalation corticosteroids* | 1 (1.1%) |
| Anosmia day 1*, n (%) |  |
| →*No* | 58 (67.4%) |
| →*Yes* | 25 (29.0%) |
| →*Unknown* | 3 (3.5%) |
| Ageusia day 1*, n (%) |  |
| →*No* | 63 (73.3%) |
| →*Yes* | 21 (24.4%) |
| →*Unknown* | 2 (2.3%) |

n: number, IQR: interquartile range, COVID-19: coronavirus disease 2019
\*not filled in by all participants

### Olfactory changes

Olfactory changes were reported by 40/94 (43%) participants at three weeks, of whom 34 (36%) had a diminished smell, seven (7%) had changes in familiar scents, four (4%) had no smell, and none experienced new unpleasant scents. Of those participants with reported olfactory changes, two had nasal congestion. At three months, reported olfactory changes were present in 17/43 (40%) participants and in 12/19 (63%) participants at six months (Fig 2A).

**Fig 2.**
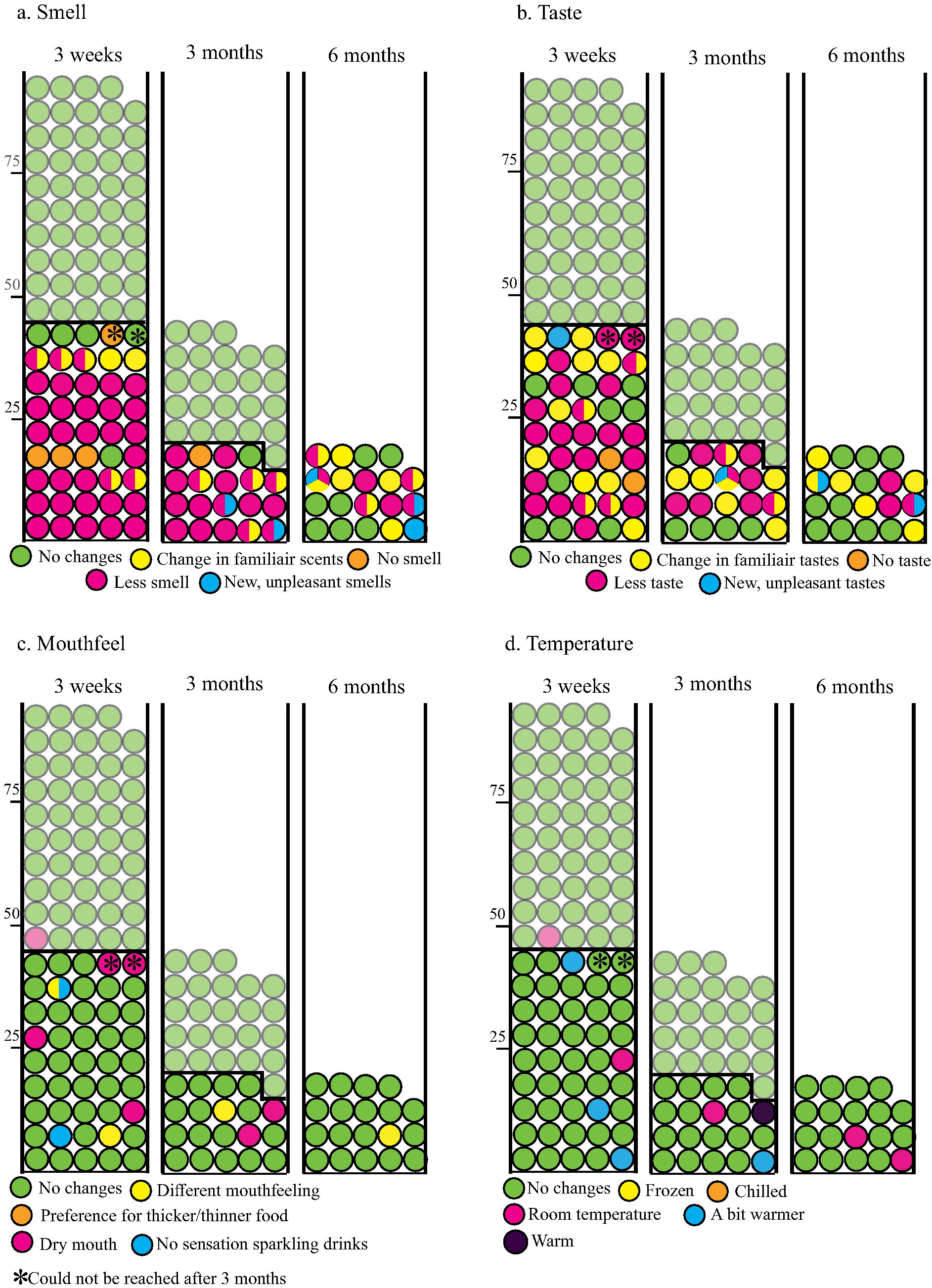
The reported type of a) smell, b) taste, c) mouthfeel, and d) temperature changes by participants at three weeks (n=94), three months (n=43), and six months (n=19). Every circle and place represents one participant. The participants above the black line (lighter green circles) reported no changes in taste or smell.

No participants reported an increase in olfactory changes at three weeks, three months and six months since being positively tested for SARS-CoV-2. The course of olfactory changes are mostly reported to be decreasing ( i.e. normalization of smell) at three weeks, and stable or fluctuating at three months and six months (Fig 3A). The degree of olfactory changes is reported to be mostly ‘a bit’ at three weeks and three months and ‘quite a bit’ at six months. Odors that were smelled more intensely were mostly smoke and feces (S2 Table).

**Fig 3.**
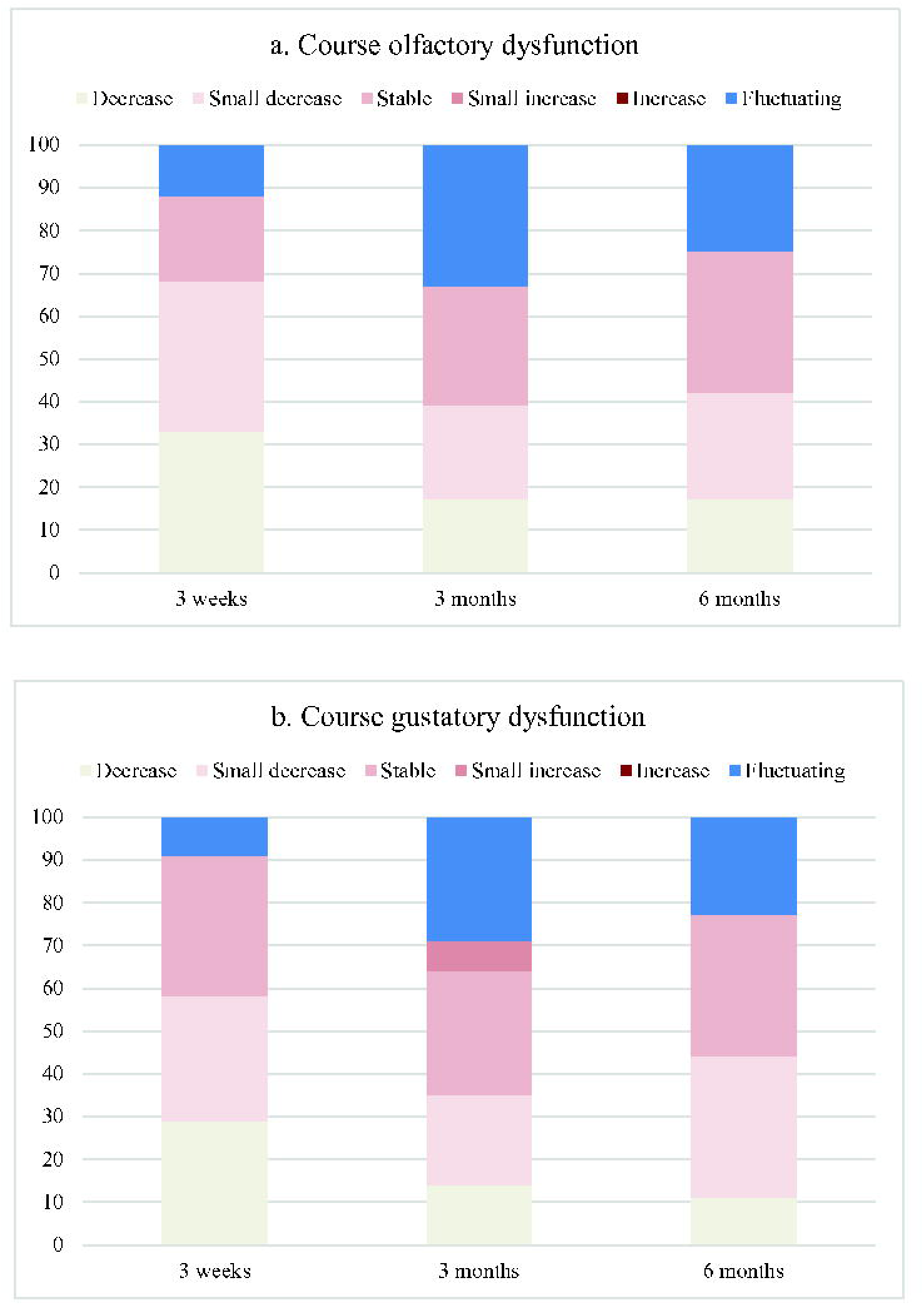
Recalled course of a) olfactory and b) gustatory changes at three weeks (n=94), three months (n=43), and six months (n=19) in percentages.

### Gustatory changes

Gustatory changes were reported by 37 of the 94 (39%) participants at three weeks; 24 (26%) tasted less than before the SARS-CoV-2 infection, 14 (15%) experienced changes in taste, two (2%) had no taste and one (1%) reported new unpleasant tastes in the mouth. At three months and six months, gustatory changes were described by 14/43 (33%) and 9/19 (47%) participants, respectively (Fig 2B).

One participant reported an increase in gustatory dysfunction at three months since being positively tested for SARS-CoV-2. Other participants reported the course of gustatory dysfunction mostly to be decreasing and stable at three weeks and six months, and stable and fluctuating at three months (Fig 3B). The degree of gustatory changes is reported to be mostly ‘a bit’ at three weeks and six months and ‘quite a bit’ at three months.

Most participants report no change or a weaker taste in the different basic tastes salt, bitter, sweet, and sour at three weeks and three months. At six months, participants mostly report no change in salt, bitter, and sour taste. However, in bitter, sweet and sour taste, participants report nearly as much a stronger and weaker intensity of these basic tastes (Fig 4A-D). A metallic and continuous taste were reported at three weeks in 5 and 9 participants, at three months in 4 and 5 participants, and at six months in 4 and 3 participants, respectively (Fig 4E-F). Participants with a continuous taste described this taste mostly as a metallic taste (4 participants at three weeks, 2 at three months, and 3 at six months). Other described tastes were something chemical, something odd, an infectious taste, bitter taste, sour taste or indescribable taste (S3 Table).

**Fig 4.**
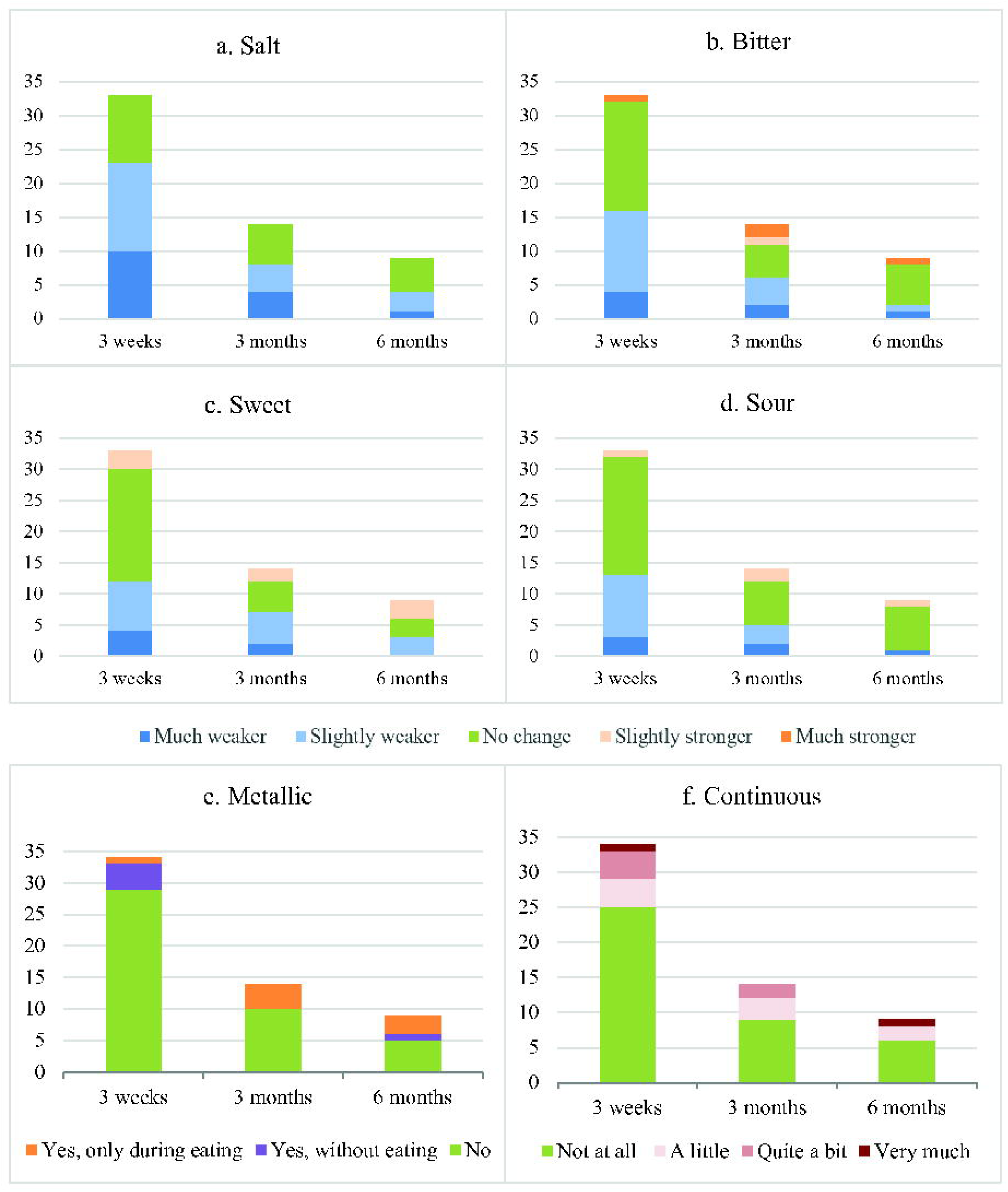
Changes in a) salt, b) bitter, c) sweet, d) sour, e) metallic, and f) continuous taste in participants with gustatory dysfunction at three weeks (n=33), three months (n=14), and six months (n=9).

### Trigeminal changes

Trigeminal changes were reported infrequently: 5/94 (5%) participants reported a dry mouth, 2/94 (2%) had a different mouthfeel, and 2/94 (2%) could not feel a tingling sensation when drinking carbonated beverages at three weeks. At three and six months, respectively 3/43 (7%) and 1/19 (5%) of the participants reported trigeminal changes (Fig 2C).

At three weeks, 2/94 (2%) and 3/94 (3%) of the participants liked the temperature of their food and drinks to be at chamber temperature and a bit warmer than normal, respectively. Changes in food temperature preference were reported by 3/43 (7%) and 2/19 (11%) participants at three months and six months, respectively (Fig 2D).

### Overview olfactory, gustatory, trigeminal changes

Most participants who report olfactory changes also report changes in taste. This overlap between the presence of smell and taste changes is observed at three weeks, three months and six months. Moreover, nearly all participants with trigeminal changes have olfactory and/or gustatory changes too. At three months and six months, trigeminal changes do not occur in isolation. Only at three months, two participants report changes in all sensory aspects (Fig 5).

**Fig 5.**
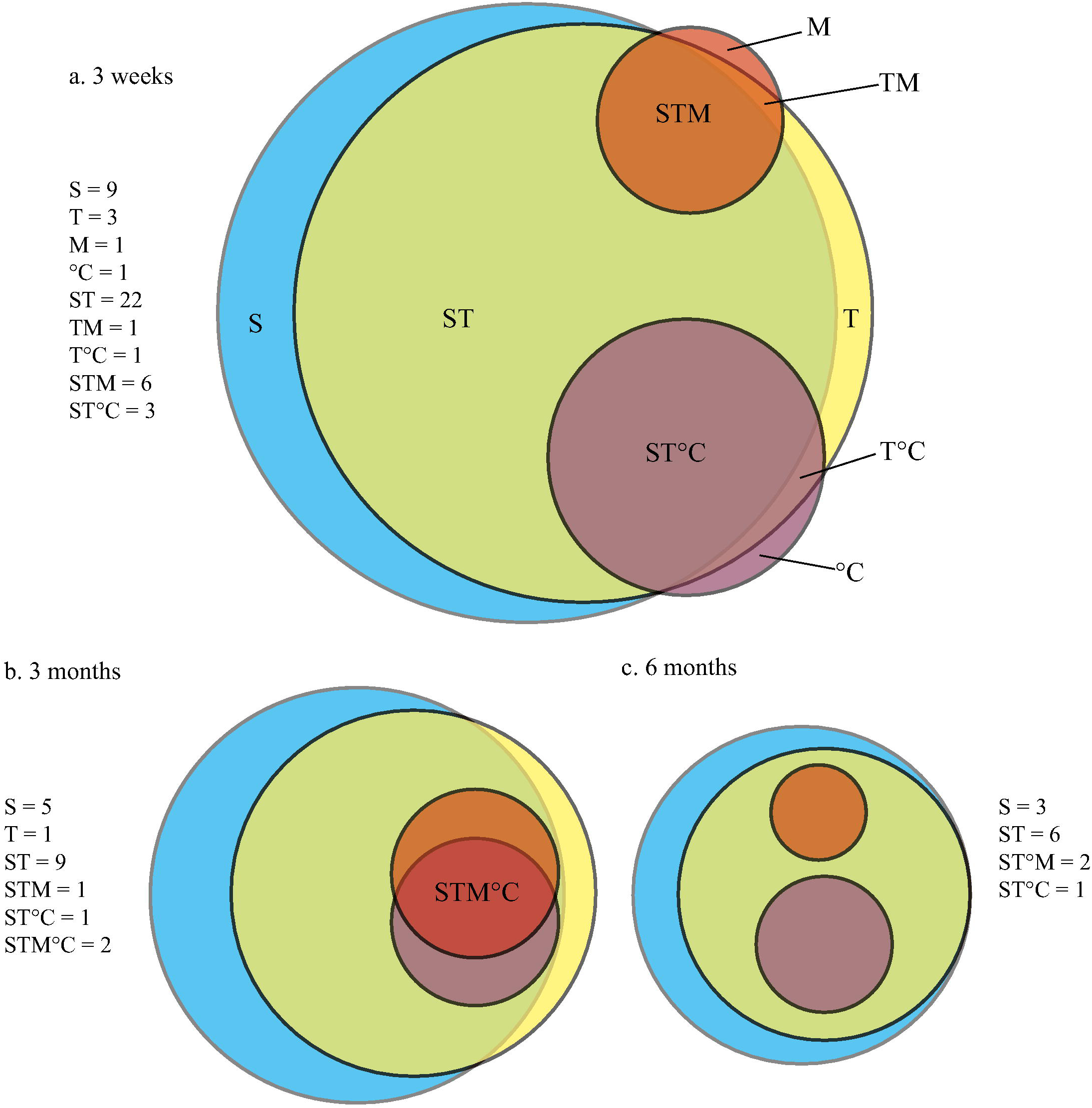
Overlap of changes in smell, taste, mouthfeel and temperature in patients with COVID-19 after a) 3 weeks, b) three months and c) 6 months. S = smell, T = taste, M = mouthfeel, °C = temperature.

### Impact on daily life and QoL of olfactory, gustatory and trigeminal changes

At three weeks, three months and six months, impact of reported olfactory changes on daily life - ranging from ‘a bit’ to ‘very much’ - is reported by 27/40 (68%), 14/18 (78%), and 10/12 (83%) participants, respectively. Most participants describe the impact on daily life as ‘a bit’ at three weeks and three months, and as ‘quite a bit’ at six months. Impact on QoL - ranging from ‘a bit’ to ‘very much’ - has been described by 14/40 (35%), 8/18 (44%) and 9/12 (75%) participants at three weeks, three months and six months, respectively. At three weeks, most participants report ‘a bit’ impact on QoL, and ‘quite a bit’ at three months and six months. (Fig 6). At three weeks 7/7 (100%) participants, at three months 4/5 (80%) participants, and at six months 6/10 (60%) participants did nothing to cope with unpleasant or changing scents. At three and at six months respectively, one and four participants avoided bad smells. Diminished smell was mostly handled by smelling things to check if they smelled anything, asking other people to smell food (for example burned food or shelf life), and paying more attention to personal hygiene. Participants also tried smell training (three at three weeks, five at three months, and two at six months) (S3 Table A-B).

**Fig 6.**
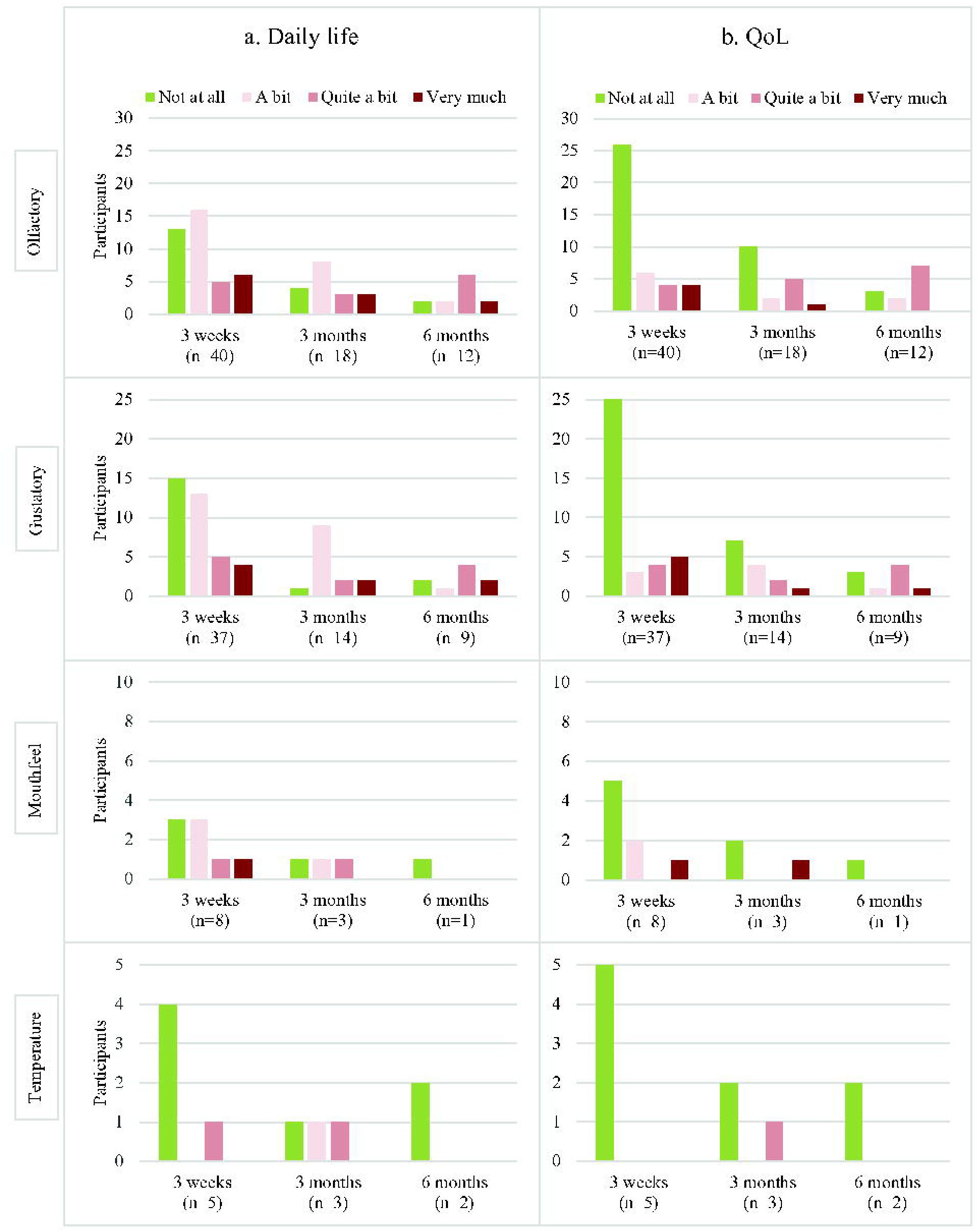
Impact on a) daily life and b) QoL of olfactory, gustatory, mouthfeel, and temperature changes.

Impact of reported gustatory changes on daily life were described by 22/37 (59%), 13/14 (93%) and 7/9 (78%) participants at three weeks, three months and six months, respectively. Most participants described the impact on daily life as ‘a bit’ at three weeks and three months, and as ‘quite a bit’ at six months. Impact of reported gustatory changes on QoL is reported by 12/37 (32%), 7/14 (50%), and 6/9 (67%) participants, respectively. At three weeks, most participants reported ‘very much’ impact on QoL, ‘a bit’ impact at three months, and ‘quite a bit’ impact at six months. (Figure 6). To cope with reported gustatory changes, at three weeks 22/37 (59%) of the participants, at three months 6/14 (43%) participants, and at six months 4/9 (44%) of the participants with these changes reported to do nothing. Some participants experimented with stronger flavors, asked for help with cooking or avoided products they did not like (S3 Table C).

Most participants reported to take no action to handle an alteration in a basic taste, although some participants avoided or added extra of a basic taste.

A metallic taste mostly had ‘a bit’ impact at three weeks (3/5, 60%) and ‘quite a bit’ at three months (2/4, 50%) and six months (2/4, 50%). To cope with a metallic taste, none of the participants reported any specific action, except for one, who avoided nutritional products with a high amount of iron. Most participants with a continuous taste reported no impact on daily life.

For reported trigeminal changes, impact of mouthfeel changes on daily life is described by 5/8, 2/3, and 0/1 participants at three weeks, three months and six months, respectively. Most participants reported ‘a bit’ impact on daily life. Impact of temperature changes is described by 1/5, 2/3, and 0/2 participants as ‘a bit’ or ‘quite a bit’ at three weeks, three months and six months, respectively. Impact on QoL has been reported by 3/8, 1/3 and 0/1 participants with mouthfeel changes as ‘a bit’ or ‘very much’, and 0/5, 1/3 and 0/2 with temperature changes as ‘quite a bit’ at three weeks, three months and six months, respectively.

At three weeks, 8/45 (18%) participants would like to receive medical guidance for their changes. However, at six months, 7/12 (58%) participants with sensory alterations would like to receive help, more specifically (experimental) medical treatment and dietary advice.

## Discussion

This study demonstrates that the prevalence of reported olfactory, gustatory and trigeminal changes is higher at three weeks after SARS-CoV-2 infection than at three and six months after SARS-CoV-2 infection in non-hospitalized patients. Furthermore, reported olfactory and gustatory changes are more present than trigeminal changes, the last playing a role only in a minority of patients. At three weeks, most patients with reported olfactory and gustatory changes characterize these changes as ‘less or no smell and taste’, respectively. Reported olfactory, gustatory and trigeminal changes have modest impact on daily life and QoL in a majority of the patients, but have a profound effect in some patients. Patients with reported sensory alterations are more interested in professional help at six months than at three weeks and three months.

Trigeminal changes were described by a minority of the patients. This is in line with other studies reporting trigeminal changes in COVID-19 patients[4,20–22]. However, previous studies did not combine longitudinal questionnaires with measurements of impact on daily life and QoL, and mostly used provocative substances which participants could sniff to study nasal trigeminal dysfunction[4,20–22]. QoL has been studied in COVID-19 patients with olfactory and gustatory changes, but not longitudinally[11,13,23]. In these previous studies and in our present study, QoL is substantially impacted by the olfactory and gustatory changes. We hypothesize that through time, patients may perceive the changes as more bothersome due to the duration, or patients can adapt to changes and perceive them as less bothersome. Therefore, the longitudinal approach in our study contributes to existing knowledge. Our study showed no difference in impact on daily life and QoL between the time points. However, the limited number of participants precludes drawing firm conclusions.

A change in familiar smells and new, unpleasant smells were, compared to less and no smells, relatively more reported at six months than at three weeks, which corresponds with a normal, physiological recovery as found in other studies[24]. Distorted smell can follow after reduced smell when new sensory pathways of an odor stimulate the wrong part of the olfactory bulb during neurogenesis[25].

This study is first to study changes in trigeminal function longitudinally in combination with impact on daily life and QoL. A strength of this study is the combination of longitudinal data on reported prevalence, characteristics and impact of olfactory, gustatory and trigeminal changes in COVID-19 patients. Both a strength and limitation of this study is the lack of objective measurement tools, such as Sniffin’ Sticks and taste strips. Subjective measurements are considered important, as it takes the patients’ experience and thereby personal bothering into account[26]. However, it should be noted that specific changes in olfactory and gustatory function are often difficult to describe and this could be underreported due to possible habituation of olfactory and gustatory changes. In general, the correlation between objective and subjective measurements is weak[26]. In this study, biases were avoided by actively calling the participants multiple times if the phone call was not answered (nonresponse bias), by maintaining broad inclusion and exclusion criteria (selection bias), and by asking the questions in a standardized and neutral format (researcher bias). The questionnaire used in this study is based on a questionnaire previously applied in patients with gastro-intestinal stromal tumors (GIST) treated with tyrosine-kinase inhibitors (TKIs)[19]. In these patients, the impact of trigeminal changes was modest too, and olfactory and gustatory changes are reported to have more impact on daily life and QoL. As the number of participants reporting sensory changes at six months is limited, the characteristics of this group could not be studied optimally. Future studies should include a larger study population to increase generalizability. Moreover, we also recommend to examine the impact of vaccination and the different variants of SARS-CoV-2 on olfactory, gustatory, and trigeminal changes, as these influence the severity and pattern of the symptoms[14,15].

In conclusion, non-hospitalized COVID-19 patients report a higher prevalence of olfactory and gustatory changes than trigeminal changes at three weeks, three months and six months. A modest impact on daily life and QoL has been reported by the participants with these changes at all time points. Health professionals should offer information, and help patients to cope with the changes.

## Supporting information

S1_Questionnaire

## Data Availability

All relevant data can be found in S1 Data.

## Acknowledgments

None

## Data Availability Statement

The datasets generated and/or analyzed during the current study are included in this published article and its supplementary material files. Additional supporting data and materials related to this study are available from the corresponding author on reasonable request.

## References

[1] J.Y. Tong, A. Wong, D. Zhu, J.H. Fastenberg, T. Tham, The Prevalence of Olfactory and Gustatory Dysfunction in COVID-19 Patients: A Systematic Review and Meta-analysis., Otolaryngol Head Neck Surg 163 (2020) 3–11. 10.1177/0194599820926473.

[2] D. Borsetto, C. Hopkins, V. Philips, R. Obholzer, G. Tirelli, J. Polesel, P. Boscolo-Rizzo, Self-reported alteration of sense of smell or taste in patients with COVID-19: a systematic review and meta-analysis on 3563 patients., Rhinology 58 (2020) 430–436. 10.4193/Rhin20.185.

[3] K. Ohla, M.G. Veldhuizen, T. Green, M.E. Hannum, A.J. Bakke, S.T. Moein, A. Tognetti, E.M. Postma, R. Pellegrino, D.L.D. Hwang, J. Albayay, S. Koyama, A.A. Nolden, T. Thomas-Danguin, C. Mucignat-Caretta, N.S. Menger, I. Croijmans, L. Öztürk, H. Yanık, D. Pierron, V. Pereda-Loth, A. Nunez-Parra, A.M. Martinez Pineda, D. Gillespie, M.C. Farruggia, C. Cecchetto, M.A. Fornazieri, C. Philpott, V. Voznessenskaya, K.W. Cooper, P. Rohlfs Dominguez, O. Calcinoni, J. de Groot, S. Boesveldt, S. Bhutani, E.M. Weir, C. Exten, P. V Joseph, J.E. Hayes, M.Y. Niv, A follow-up on quantitative and qualitative olfactory dysfunction and other symptoms in patients recovering from COVID-19 smell loss., Rhinology (2022). 10.4193/Rhin21.415.

[4] V. Parma, K. Ohla, M.G. Veldhuizen, M.Y. Niv, C.E. Kelly, A.J. Bakke, K.W. Cooper, C. Bouysset, N. Pirastu, M. Dibattista, R. Kaur, M.T. Liuzza, M.Y. Pepino, V. Schöpf, V. Pereda-Loth, S.B. Olsson, R.C. Gerkin, P. Rohlfs Domínguez, J. Albayay, M.C. Farruggia, S. Bhutani, A.W. Fjaeldstad, R. Kumar, A. Menini, M. Bensafi, M. Sandell, I. Konstantinidis, A. Di Pizio, F. Genovese, L. Öztürk, T. Thomas-Danguin, J. Frasnelli, S. Boesveldt, Ö. Saatci, L.R. Saraiva, C. Lin, J. Golebiowski, L.-Dar Hwang, M.H. Ozdener, M.D. Guàrdia, C. Laudamiel, M. Ritchie, J. Havlícek, D. Pierron, E. Roura, M. Navarro, A.A. Nolden, J. Lim, K.L. Whitcroft, L.R. Colquitt, C. Ferdenzi, E. V Brindha, A. Altundag, A. Macchi, A. Nunez-Parra, Z.M. Patel, S. Fiorucci, C.M. Philpott, B.C. Smith, J.N. Lundström, C. Mucignat, J.K. Parker, M. van den Brink, M. Schmuker, F.P.S. Fischmeister, T. Heinbockel, V.D.C. Shields, F. Faraji, E. Santamaría, W.E.A. Fredborg, G. Morini, J.K. Olofsson, M. Jalessi, N. Karni, A. D’Errico, R. Alizadeh, R. Pellegrino, P. Meyer, C. Huart, B. Chen, G.M. Soler, M.K. Alwashahi, A. Welge-Lüssen, J. Freiherr, J.H.B. de Groot, H. Klein, M. Okamoto, P.B. Singh, J.W. Hsieh, D.R. Reed, T. Hummel, S.D. Munger, J.E. Hayes, More than smell - COVID-19 is associated with severe impairment of smell, taste, and chemesthesis., Chem Senses (2020). 10.1093/chemse/bjaa041.

[5] M.V. Cuevas-Gonzalez, L.F. Espinosa-Cristóbal, A. Donohue-Cornejo, K.L. Tovar-Carrillo, R.A. Saucedo-Acuña, A.G. García-Calderón, D.A. Guzmán-Gastelum, J.C. Cuevas-Gonzalez, COVID-19 and its manifestations in the oral cavity: A systematic review., Medicine 100 (2021) e28327. 10.1097/MD.0000000000028327.

[6] L.M. Bartoshuk, G.K. Beauchamp, Chemical senses., Annu Rev Psychol 45 (1994) 419–449. 10.1146/annurev.ps.45.020194.002223.

[7] R.E.A. Santos, M.G. da Silva, M.C.B. do Monte Silva, D.A.M. Barbosa, A.L. do V. Gomes, L.C.M. Galindo, R. da Silva Aragão, K.N. Ferraz-Pereira, Onset and duration of symptoms of loss of smell/taste in patients with COVID-19: A systematic review., Am J Otolaryngol 42 (2021) 102889. 10.1016/j.amjoto.2020.102889.

[8] P. Boscolo-Rizzo, F. Guida, J. Polesel, A.V. Marcuzzo, P. Antonucci, V. Capriotti, E. Sacchet, F. Cragnolini, A. D’Alessandro, E. Zanelli, R. Marzolino, C. Lazzarin, M. Tofanelli, N. Gardenal, D. Borsetto, C. Hopkins, L.A. Vaira, G. Tirelli, Self-reported smell and taste recovery in coronavirus disease 2019 patients: a one-year prospective study., Eur Arch Otorhinolaryngol 279 (2022) 515–520. 10.1007/s00405-021-06839-w.

[9] A. Pavli, M. Theodoridou, H.C. Maltezou, Post-COVID Syndrome: Incidence, Clinical Spectrum, and Challenges for Primary Healthcare Professionals., Arch Med Res 52 (2021) 575–581. 10.1016/j.arcmed.2021.03.010.

[10] T. Miwa, M. Furukawa, T. Tsukatani, R.M. Costanzo, L.J. DiNardo, E.R. Reiter, Impact of olfactory impairment on quality of life and disability., Arch Otolaryngol Head Neck Surg 127 (2001) 497–503. 10.1001/archotol.127.5.497.

[11] D.H. Coelho, E.R. Reiter, S.G. Budd, Y. Shin, Z.A. Kons, R.M. Costanzo, Quality of life and safety impact of COVID-19 associated smell and taste disturbances., Am J Otolaryngol 42 (2021) 103001. 10.1016/j.amjoto.2021.103001.

[12] T. Hummel, S. Nordin, Olfactory disorders and their consequences for quality of life., Acta Otolaryngol 125 (2005) 116–121. 10.1080/00016480410022787.

[13] S.M.A. Elkholi, M.K. Abdelwahab, M. Abdelhafeez, Impact of the smell loss on the quality of life and adopted coping strategies in COVID-19 patients., Eur Arch Otorhinolaryngol (2021) 1–8. 10.1007/s00405-020-06575-7.

[14] P. Boscolo-Rizzo, G. Tirelli, P. Meloni, C. Hopkins, G. Madeddu, A. De Vito, N. Gardenal, R. Valentinotti, M. Tofanelli, D. Borsetto, J.R. Lechien, J. Polesel, G. De Riu, L.A. Vaira, Coronavirus disease 2019 (COVID-19)-related smell and taste impairment with widespread diffusion of severe acute respiratory syndrome-coronavirus-2 (SARS-CoV-2) Omicron variant., Int Forum Allergy Rhinol (2022). 10.1002/alr.22995.

[15] L.A. Vaira, A. De Vito, J.R. Lechien, C.M. Chiesa-Estomba, M. Mayo-Yàñez, C. Calvo-Henrìquez, S. Saussez, G. Madeddu, S. Babudieri, P. Boscolo-Rizzo, C. Hopkins, G. De Riu, New Onset of Smell and Taste Loss Are Common Findings Also in Patients With Symptomatic COVID-19 After Complete Vaccination., Laryngoscope 132 (2022) 419–421. 10.1002/lary.29964.

[16] A. Tami, B.T.F. van der Gun, K.I. Wold, M.F. Vincenti-González, A.C.M. Veloo, M. Knoester, V.P.R. Harmsma, G.C. de Boer, A.L.W. Huckriede, D. Pantano, L. Gard, I.A. Rodenhuis-Zybert, V. Upasani, J. Smit, A.E. Dijkstra, J.J. de Haan, J.M. van Elst, J. van den Boogaard, S. O’ Boyle, L. Nacul, H.G.M. Niesters, A.W. Friedrich, The COVID HOME study research protocol: Prospective cohort study of non-hospitalised COVID-19 patients, PLoS One 17 (2022) e0273599-. 10.1371/journal.pone.0273599.

[17] I. IJpma, E.R. Timmermans, R.J. Renken, G.J. Ter Horst, A.K.L. Reyners, Metallic Taste in Cancer Patients Treated with Systemic Therapy: A Questionnaire-based Study., Nutr Cancer 69 (2017) 140–145. 10.1080/01635581.2017.1250922.

[18] J.M. van Elst, S. Boesveldt, A. Vissink, H. Jager-Wittenaar, A.K.L. Reyners, J.J. de Haan, Subjective Mouthfeel and Temperature Alterations in COVID-19 Patients Six to Ten Months After Diagnosis, Chemosens Percept 15 (2022) 165–174. 10.1007/s12078-022-09304-y.

[19] J.M. van Elst, N.S. IJzerman, R.H.J. Mathijssen, N. Steeghs, A.K.L. Reyners, J.J. de Haan, Taste, smell and mouthfeel disturbances in patients with gastrointestinal stromal tumors treated with tyrosine-kinase inhibitors., Support Care Cancer 30 (2022) 2307– 2315. 10.1007/s00520-021-06658-z.

[20] P. Boscolo-Rizzo, T. Hummel, C. Hopkins, M. Dibattista, A. Menini, G. Spinato, C. Fabbris, E. Emanuelli, A. D’Alessandro, R. Marzolino, E. Zanelli, E. Cancellieri, K. Cargnelutti, S. Fadda, D. Borsetto, L.A. Vaira, N. Gardenal, J. Polesel, G. Tirelli, High prevalence of long-term olfactory, gustatory, and chemesthesis dysfunction in post-COVID-19 patients: a matched case-control study with one-year follow-up using a comprehensive psychophysical evaluation., Rhinology 59 (2021) 517–527. 10.4193/Rhin21.249.

[21] B. Iravani, A. Arshamian, A. Ravia, E. Mishor, K. Snitz, S. Shushan, Y. Roth, O. Perl, D. Honigstein, R. Weissgross, S. Karagach, G. Ernst, M. Okamoto, Z. Mainen, E. Monteleone, C. Dinnella, S. Spinelli, F. Mariño-Sánchez, C. Ferdenzi, M. Smeets, K. Touhara, M. Bensafi, T. Hummel, N. Sobel, J.N. Lundström, Relationship between odor intensity estimates and COVID-19 prevalence prediction in a Swedish population., Chem Senses (2020). 10.1093/chemse/bjaa034.

[22] F. Ferreli, M. Di Bari, F. Gaino, A. Albanese, L.S. Politi, G. Spriano, G. Mercante, Trigeminal features in COVID-19 patients with smell impairment., Int Forum Allergy Rhinol 11 (2021) 1253–1255. 10.1002/alr.22796.

[23] M.S. Otte, A. Haehner, M.-L. Bork, J.P. Klussmann, J.C. Luers, T. Hummel, Impact of COVID-19-Mediated Olfactory Loss on Quality of Life., ORL J Otorhinolaryngol Relat Spec (2022) 1–6. 10.1159/000523893.

[24] B. Prem, D.T. Liu, G. Besser, G. Sharma, L.E. Dultinger, S. V Hofer, M.M. Matiasczyk, B. Renner, C.A. Mueller, Long-lasting olfactory dysfunction in COVID-19 patients., Eur Arch Otorhinolaryngol 279 (2022) 3485–3492. 10.1007/s00405-021-07153-1.

[25] R. Pellegrino, J.D. Mainland, C.E. Kelly, J.K. Parker, T. Hummel, Prevalence and correlates of parosmia and phantosmia among smell disorders, Chem Senses 46 (2021) bjab046. 10.1093/chemse/bjab046.

[26] L.E. Spotten, C.A. Corish, C.M. Lorton, P.M. Ui Dhuibhir, N.C. O’Donoghue, B. O’Connor, T.D. Walsh, Subjective and objective taste and smell changes in cancer., Ann Oncol 28 (2017) 969–984. 10.1093/annonc/mdx018.

