## Supplementary materials for "Olfactory, gustatory and trigeminal changes in non-hospitalized COVID-19 patients: an exploratory prospective cohort study"

**S1_Questionnaire. Questionnaire on sensory changes**

**S1_Data. All relevant data.**

**S1_Table. Odors perceived more sensitively by participants with olfactory changes.**

**S2_Table. Coping with smell and taste changes.**

**S3_Table. Resemblance continuous taste.**
