## Supplementary material for "Olfactory, gustatory and trigeminal changes in non-hospitalized COVID-19 patients: an exploratory prospective cohort study": S1_Questionnaire

**S1 Questionnaire. Questionnaire on sensory changes**

**Smell, taste, texture and temperature changes**

| Q1a. | How has your **sense of smell** changed compared to before being positively tested for the corona virus, even if just a little?    **CIRCLE ONE CODE ONLY** | | |
| --- | --- | --- | --- |
| I do not perceive any smell at all | | 1 | **CONTINUE TO Q1b** |
| I perceive a difference in intensity of smells in general/lack of smell (smell acuity) | | 2 |  |
| I don’t perceive food smells in the same way I did before (smell alteration) | | 3 |  |
| I detect new bad smells | | 4 |  |
| I do not perceive any change in taste perception | | 5 | **SKIP TO Q2a** |

| Q1b. | | **ONLY ASK THOSE WHO SELECTED 1-4 IN Q1a**  Thinking about the things that have changed your **sense of smell**, how much does it impact your daily life?  **CIRCLE ONE CODE ONLY** | | | | |
| --- | --- | --- | --- | --- | --- | --- |
|  | | | Not at all | A little | Quite a bit | Very much |
|  | Smell | | 1 | 2 | 3 | 4 |

| Q1c. | | Thinking about the things that have changed your **sense of smell,** is your quality of life negatively affected because of these changes in taste?  **CIRCLE ONE CODE ONLY** | | | | |
| --- | --- | --- | --- | --- | --- | --- |
|  | | | Not at all | A little | Quite a bit | Very much |
|  | Smell | | 1 | 2 | 3 | 4 |

| Q2a. | How has your **sense of taste** changed compared to before being positively tested for the corona virus, even if just a little?  **CIRCLE ALL THAT APPLY** | | |
| --- | --- | --- | --- |
| I do not perceive any taste anymore | | 1 | **CONTINUE TO Q2b** |
| I perceive a difference in intensity of tastes in general  (lack of taste) | | 2 |  |
| I don’t perceive certain tastes the same way I did before  (taste alteration) | | 3 |  |
| I detect new bad tastes in my mouth | | 4 |  |
| I do not perceive any change in taste perception | | 5 | **SKIP TO Q3a** |

| Q2b. | | **ONLY ASK THOSE WHO SELECTED 1-4 IN Q2a**  Thinking about the things that have changed your **sense of taste** (e.g. sweet, salt, sour, bitter, metallic taste), how much does it impact your daily life?  **CIRCLE ONE CODE ONLY** | | | | |
| --- | --- | --- | --- | --- | --- | --- |
|  | | | Not at all | A little | Quite a bit | Very much |
|  | Taste | | 1 | 2 | 3 | 4 |

| Q2c. | | Thinking about the things that have changed your **sense of taste** (e.g. sweet, salt, sour, bitter, metallic taste), is your quality of life negatively affected because of these changes in taste?  **CIRCLE ONE CODE ONLY** | | | | |
| --- | --- | --- | --- | --- | --- | --- |
|  | | | Not at all | A little | Quite a bit | Very much |
|  | Taste | | 1 | 2 | 3 | 4 |

| Q3a. | How has your **sense of mouthfeel**, changed compared to before being positively tested for the corona virus, even if just a little?    **CIRCLE ALL THAT APPLY** | | |
| --- | --- | --- | --- |
| I perceive mouthfeel different than before | | 1 | **CONTINUE TO Q3b** |
| I do perceive a dry mouth | | 2 |  |
| I prefer other textures (e.g. thicker/thinner) than before | | 3 |  |
| I don’t feel the tingling sensation of carbonated drinks anymore | | 4 |  |
| I do not perceive any change in mouthfeel perception | | 5 | **SKIP TO Q4a** |

| Q3b. | | **ONLY ASK THOSE WHO SELECTED 1-4 IN Q3a**  Thinking about the things that have changed your **sense of mouthfeel** (e.g. dry mouth feelings, temperature perception), how much does it impact your daily life?  **CIRCLE ONE CODE ONLY** | | | | |
| --- | --- | --- | --- | --- | --- | --- |
|  | | | Not at all | A little | Quite a bit | Very much |
|  | Mouthfeel | | 1 | 2 | 3 | 4 |

| Q3c. | | Thinking about the things that have changed your **sense of mouthfeel** (e.g. dry mouth feelings, temperature perception)**,** is your quality of life negatively affected because of these changes in taste?  **CIRCLE ONE CODE ONLY** | | | | |
| --- | --- | --- | --- | --- | --- | --- |
|  | | | Not at all | A little | Quite a bit | Very much |
|  | Mouthfeel | | 1 | 2 | 3 | 4 |

| Q4a. | Do you have a change in preference on the **specific temperature at which food and drinks are served,** compared to before being positively tested for the corona virus**,** even if just a little?    **CIRCLE ALL THAT APPLY** | | |
| --- | --- | --- | --- |
| Yes, I prefer frozen now | | 1 | **CONTINUE TO Q4b** |
| Yes, I prefer chilled (fridge temperature) now | | 2 |  |
| Yes, I prefer ambient temperature now (around 20 degree Celsius) | | 3 |  |
| Yes, I prefer a little warmer now | | 4 |  |
| Yes, I prefer hot now | | 5 |  |
| No, it remains the same | | 6 |  |

| Q4b. | | **ONLY ASK THOSE WHO SELECTED 1-4 IN Q4a**  Thinking about the things that have changed your **specific temperature at which food and drinks are served,** how much does it impact your daily life?  **CIRCLE ONE CODE ONLY** | | | | |
| --- | --- | --- | --- | --- | --- | --- |
|  | | | Not at all | A little | Quite a bit | Very much |
|  | Temperature | | 1 | 2 | 3 | 4 |

| Q4c. | | Thinking about the things that have changed your **specific temperature at which food and drinks are served,** is your quality of life negatively affected because of these changes in taste?  **CIRCLE ONE CODE ONLY** | | | | |
| --- | --- | --- | --- | --- | --- | --- |
|  | | | Not at all | A little | Quite a bit | Very much |
|  | Temperature | | 1 | 2 | 3 | 4 |

- **IF RESPONDENT HAS SELECTED CODES 1-4 AT Q1a, ASK QUESTION Q6a.**
- **IF RESPONDENT HAS SELECTED CODE 5 AT Q1a AND IF RESPONDENT HAS SELECTED CODES 1-4 AT Q2a, ASK QUESTION Q6a.**
- **IF RESPONDENT HAS SELECTED CODE 5 AT Q1a AND Q2a, THE QUESTIONNAIRE IS COMPLETED.**

**SMELL**

**RESEARCHER: ASK QUESTION Q5a IF RESPONDENT HAS SELECTED CODES 1-4 AT Q1a (SMELL PERCEPTIONS HAVE CHANGED).**

| Q5a. | | Thinking about your experiences of eating and drinking **since you have been tested positive for the corona virus**, to what level of severity did your smell perception change?  **CIRCLE ONE CODE ONLY** | | | | |
| --- | --- | --- | --- | --- | --- | --- |
|  | | | Not at all | A little | Quite a bit | Very much |
| a. | Smell | | 1 | 2 | 3 | 4 |
|  |  | |  | **CONTINUE TO Q5b** | | |

| Q5b. | | **ONLY ASK THOSE WHO SELECTED 2-4 IN Q5a**  How is the **course** of the smell changes since you have been tested positive for the corona virus?  **CIRCLE ONE CODE ONLY** | | | | | | |
| --- | --- | --- | --- | --- | --- | --- | --- | --- |
|  | | | It decreases / I’m almost back at my old level. | It decreases a little | Stable | Increases a little | Increases | Fluctuating |
|  | Course | | 1 | 2 | 3 | 4 | 5 | 6 |

| Q5c. | Do you have continuous nasal congestion since you have been tested positive for the corona virus?  **CIRCLE ONE CODE ONLY** | |
| --- | --- | --- |
| No | | 1 |
| At the start, but not anymore | | 2 |
| Yes | | 3 |

| Q5d. | **ASK AS OPEN QUESTION**  Generally, what do you do to cope with bad or distorted smells?  **CIRCLE ALL THAT APPLY** | |
| --- | --- | --- |
| Nothing | | 1 |
| I choose colder foods (sandwiches, salads, fruits, and cheeses) that are less aromatic | | 2 |
| I open the windows and use the exhaust fan when cooking; open containers away from the face | | 3 |
| I eat other types of food that have smells I can bear | | 4 |
| Other, please specify:_______________ | | 5 |

| Q5e. | **ASK AS OPEN QUESTION**  Generally, what do you do to cope with diminished smell?  **CIRCLE ALL THAT APPLY** | |
| --- | --- | --- |
| I don’t have a diminished smell | | 1 |
| Nothing | | 2 |
| I check more often if the gas is turned off | | 3 |
| I smell at products to check if I do smell something | | 4 |
| I ask other people to smell at possible rotten foods | | 5 |
| Other, please specify:_______________ | | 6 |

| Q5f. | **ASK AS OPEN QUESTION**  Can you please specify what, if any, odors you feel more sensitively than before?  **CIRCLE ALL THAT APPLY** | |
| --- | --- | --- |
| Cleaning products | | 1 |
| Perfume | | 2 |
| Odor of the hospital room | | 3 |
| Fish | | 4 |
| Dairy (milk) | | 5 |
| Meat | | 6 |
| Acidic fruit | | 7 |
| Sweet fruits | | 8 |
| Savory | | 9 |
| Food cooking | | 10 |
| Other, please specify:_____ | | 11 |
| None | | 12 |

- **IF RESPONDENT HAS SELECTED CODES 1-4 AT Q2a, ASK QUESTION Q6a.**
- **IF RESPONDENT HAS SELECTED CODE 5 AT Q2a, ASK QUESTION Q13**

**TASTE**

**RESEARCHER: ASK QUESTION Q6a IF RESPONDENT HAS SELECTED CODES 1-4 AT Q2a (TASTE PERCEPTIONS HAVE CHANGED).**

| Q6a. | | Thinking about your experiences of eating and drinking **since you have been tested positive for the corona virus**, to what level of severity did your taste perception change?  **CIRCLE ONE CODE ONLY** | | | | |
| --- | --- | --- | --- | --- | --- | --- |
|  | | | Not at all | A little | Quite a bit | Very much |
| a. | Taste  (e.g. sweet, salty, sour, bitter, metallic etc.) | | 1 | 2 | 3 | 4 |
|  |  | | **SKIP TO Q13** | **CONTINUE TO Q6b** | | |

| Q6b. | | **ONLY ASK THOSE WHO SELECTED 2-4 IN Q6a.**  How is the **course** of the taste changes since you have been tested positive for the corona virus?  **CIRCLE ONE CODE ONLY** | | | | | | |
| --- | --- | --- | --- | --- | --- | --- | --- | --- |
|  | | | It decreases / I’m almost back at my old level. | It decreases a little | Stable | Increases a little | Increases | Fluctuating |
|  | Course | | 1 | 2 | 3 | 4 | 5 | 6 |

| Q6c. | **ASK AS OPEN QUESTION.**  Generally, what do you do to cope with the **change in taste** **intensity**?  **CIRCLE ALL THAT APPLY** | |
| --- | --- | --- |
| Nothing | | 1 |
| Experiment with stronger flavors such as spices and marinades | | 2 |
| Avoid stronger flavors, like not adding spices or pepper | | 3 |
| Choose soft foods such as baked chicken, potatoes, paste and rice | | 4 |
| Consume foods with high moisture or water content | | 5 |
| Add sauces or gravy to moisten foods | | 6 |
| Other, please specify:................................ | | 7 |

| Q7a. | | We would now like to ask you a bit more specifically about certain defined tastes. To what extent has the following **taste** **changed in intensity** for you?  **CIRCLE ONE CODE ONLY** | | | | | |
| --- | --- | --- | --- | --- | --- | --- | --- |
|  | | | Much stronger | Slightly stronger | No change | Slightly weaker | Much weaker / cannot taste it at all |
|  | A change in things that normally would taste **salty** | | 1 | 2 | 3 | 4 | 5 |
|  |  | | **CONTINUE TO Q7b** | | **SKIP TO Q8a** | **CONTINUE TO Q7b** | |

| Q7b. | | **ONLY ASK THOSE WHO SELECTED CODES 1, 2, 4 OR 5 at Q7a**  How much does the **change in** **salty taste** impact you?  **CIRCLE ONE CODE ONLY** | | | | |
| --- | --- | --- | --- | --- | --- | --- |
|  | | | Not at all | A little | Quite a bit | Very much |
|  | **Salty taste** | | 1 | 2 | 3 | 4 |
|  |  | | **SKIP TO Q8a** | **CONTINUE TO Q7c** | | |

| Q7c. | **ASK AS OPEN QUESTION**  **ONLY ASK THOSE WHO SELECTED CODES 2-4 at Q8b**  What do you do to cope with this change in **salty taste**?  **CIRCLE ALL THAT APPLY** | |
| --- | --- | --- |
| Nothing | | 1 |
| I limit salty foods and foods with added salt | | 2 |
| I do not add salt during the cooking process or after meal preparation | | 3 |
| I add fish sauce or soy sauce to the food | | 4 |
| I add additional salt to the food | | 5 |
| I add more chicken essence / chicken powder to the food | | 6 |
| Other, please specify:_____________ | | 7 |

| Q8a. | | To what extent has the following taste changed in intensity for you?  **CIRCLE ONE CODE ONLY** | | | | | |
| --- | --- | --- | --- | --- | --- | --- | --- |
|  | | | Much stronger | Slightly stronger | No change | Slightly weaker | Much weaker / cannot taste it at all |
|  | A change in things that normally would taste **bitter** | | 1 | 2 | 3 | 4 | 5 |
|  |  | | **CONTINUE TO Q8b** | | **SKIP TO Q9a** | **CONTINUE TO Q8b** | |

| Q8b. | | **ONLY ASK THOSE WHO SELECTED CODES 1, 2, 4 OR 5 at Q8a**  How much does the **change in** **bitter taste** impact you?  **CIRCLE ONE CODE ONLY** | | | | |
| --- | --- | --- | --- | --- | --- | --- |
|  | | | Not at all | A little | Quite a bit | Very much |
|  | **Bitter taste** | | 1 | 2 | 3 | 4 |
|  |  | | **SKIP TO Q9a** | **CONTINUE TO Q8c** | | |

| Q8c. | **ASK AS OPEN QUESTION**  **ONLY ASK THOSE WHO SELECTED CODES 2-4 at Q8b**  What do you do to cope with this change in **bitter taste**?  **CIRCLE ALL THAT APPLY** | |
| --- | --- | --- |
| Nothing | | 1 |
| I eat room-temperature or cold foods | | 2 |
| I limit intake of bitter products | | 3 |
| I try to make my food more sour (adding lime, lemon, vinegar or other) | | 4 |
| Other, please specify:_____________ | | 5 |

| Q9a. | | To what extent has the following taste changed in intensity for you?  **CIRCLE ONE CODE ONLY** | | | | | |
| --- | --- | --- | --- | --- | --- | --- | --- |
|  | | | Much stronger | Slightly stronger | No change | Slightly weaker | Much weaker / cannot taste it at all |
|  | A change in things that normally would taste **sweet.** | | 1 | 2 | 3 | 4 | 5 |
|  |  | | **CONTINUE TO Q9b** | | **SKIP TO Q10a** | **CONTINUE TO Q9b** | |

| Q9b. | | **ONLY ASK THOSE WHO SELECTED CODES 1, 2, 4 OR 5 at Q9a**  How much does the **change in** **sweet taste** impact you?  **CIRCLE ONE CODE ONLY** | | | | |
| --- | --- | --- | --- | --- | --- | --- |
|  | | | Not at all | A little | Quite a bit | Very much |
|  | **Sweet taste** | | 1 | 2 | 3 | 4 |
|  |  | | **SKIP TO Q10a** | **CONTINUE TO Q9c** | | |

| Q9c. | **ASK AS OPEN QUESTION**  **ONLY ASK THOSE WHO SELECTED CODES 2-4 at Q10B.**  What do you do to cope with this change in sweet taste?  **CIRCLE ONE CODE ONLY** | |
| --- | --- | --- |
| Nothing | | 1 |
| I limit sweet foods and foods with added sugar | | 2 |
| I add salt to sweet food | | 3 |
| I add more water to make the food less sweet | | 4 |
| I add sugar into the food | | 5 |
| I add sweeteners into the food such as strawberries, vanilla, etc. | | 6 |
| Other, please specify:_____________ | | 7 |

| Q10a. | | To what extent has the following taste changed in intensity for you?  **CIRCLE ONE CODE ONLY** | | | | | |
| --- | --- | --- | --- | --- | --- | --- | --- |
|  | | | Much stronger | Slightly stronger | No change | Slightly weaker | Much weaker / cannot taste it at all |
|  | A change in things that normally would taste **sour.** | | 1 | 2 | 3 | 4 | 5 |
|  |  | | **CONTINUE TO Q10b** | | **SKIP TO Q11a** | **CONTINUE TO Q10b** | |

| Q10b. | | **ONLY ASK THOSE WHO SELECTED CODES 1, 2, 4 OR 5 at Q10a**  How much does the **change in** **sour taste** impact you?  **CIRCLE ONE CODE ONLY** | | | | |
| --- | --- | --- | --- | --- | --- | --- |
|  | | | Not at all | A little | Quite a bit | Very much |
|  | **Sour taste** | | 1 | 2 | 3 | 4 |
|  |  | | **SKIP TO Q11a** | **CONTINUE TO Q10c** | | |

| Q10c. | **ASK AS OPEN QUESTION**  **ONLY ASK THOSE WHO SELECTED 2-4 AT VRAAG Q11b.**  What do you do to cope with this change in sour taste?  **CIRCLE ALL THAT APPLY** | |
| --- | --- | --- |
| Nothing | | 1 |
| I limit sour foods such as citrus fruits. | | 2 |
| I add sugar into the food to make it less sour. | | 3 |
| I try to make the food more sour (adding lime, lemon, vinegar) | | 4 |
| Other, please specify:__________________________ | | 5 |

| Q11a. | Do you experience any **metallic tastes** (e.g. imagine if you bite a key / smell your finger after you hold a key for a while) compared to before infection with the corona virus?    **CIRCLE ONE CODE ONLY** | | |
| --- | --- | --- | --- |
| No | | 1 | **SKIP TO Q11a** |
| Yes, I experience it without being presented with a food product | | 2 | **CONTINUE TO Q10b** |
| Yes, I experience it only when being presented with a food product | | 3 |  |

| Q11b. | | **ONLY ASK THOSE WHO SELECTED 2-3 IN Q11a**  Thinking about the things that make you experience any **metallic tastes,** how much does it impact your daily life?  **CIRCLE ONE CODE ONLY** | | | | |
| --- | --- | --- | --- | --- | --- | --- |
|  | | | Not at all | A little | Quite a bit | Very much |
|  | Metallic tastes | | 1 | 2 | 3 | 4 |
|  |  | | **SKIP TO Q12a** | **CONTINUE TO Q11c** | | |

| Q11c. | **ASK AS OPEN QUESTION**  **ONLY ASK THOSE WHO SELECTED CODES 2-4 at Q11b**  What do you do to cope with this change in **metallic sensation**?  **CIRCLE ALL THAT APPLY** | |
| --- | --- | --- |
| Nothing | | 1 |
| I limit red meat, liver, intestine, and other foods high in iron | | 2 |
| I experiment with other high-protein food choices such as egg, poultry, fish, beans and soy | | 3 |
| I use plastic utensils instead of metal | | 4 |
| I avoid consuming foods from metal cans and/or avoid cooking in an iron skillet | | 5 |
| Other, please specify:__________________ | | 6 |

| Q12a. |  | Patients may experience a continuous taste in their mouth. Have you experienced a **continuous taste** in your mouth that you did not experience before your infection with the corona virus?  **CIRCLE ONE CODE ONLY** | | | |
| --- | --- | --- | --- | --- | --- |
|  | | Not at all | A little | Quite a bit | Very much |
| Continuous taste | | 1 | 2 | 3 | 4 |
|  | |  | **CONTINUE TO Q12b** | | |

| Q12b. | | **ONLY ASK THOSE WHO SELECTED 2-4 IN Q12a**  Thinking about the things that make you experience any **continuous taste,** how much does it impact your daily life?  **CIRCLE ONE CODE ONLY** | | | | |
| --- | --- | --- | --- | --- | --- | --- |
|  | | | Not at all | A little | Quite a bit | Very much |
|  | Continuous taste | | 1 | 2 | 3 | 4 |

| Q12c. | | **ASK AS OPEN QUESTION**  Thinking about the things that make you experience any **continuous tastes,** which resembles this taste?  **CIRCLE ALL THAT APPLY** |
| --- | --- | --- |
| 1 | Blood | |
| 2 | Bitter | |
| 3 | Something chemical | |
| 4 | Something musty | |
| 5 | Drugs | |
| 6 | Metallic | |
| 7 | Sweet | |
| 8 | Salty | |
| 9 | Sour | |
| 10 | Other, please specify____________________________________________________________ | |

| Q13. | **ASK AS OPEN QUESTION**  **ONLY ASK THOSE WHO SELECTED 1-4 IN Q1a and/or Q2a**  Would you like more guidance for your changes in smell of taste? If yes, what kind of guidance?  **CIRCLE ALL THAT APPLY** | |
| --- | --- | --- |
| No | | 1 |
| I would like a medicament (experimental) treatment | | 2 |
| I would like to receive more information about smell and taste changes | | 3 |
| I would like to receive help from a dietitian to advise me on adapting my food to my changes in smell and taste | | 4 |
| I would like to have my family receiving more information about my smell and taste changes | | 5 |
| Other, please specify:________________________ | | 6 |
