## Supplementary material for "Olfactory, gustatory and trigeminal changes in non-hospitalized COVID-19 patients: an exploratory prospective cohort study": S1_Table

**S1 Table. Odors perceived more sensitively by participants with olfactory changes.**

|  | **3 weeks** (n=40) | **3 months**  (n=18) | **6 months**  (n=12) |
| --- | --- | --- | --- |
| Smoke | 2 | 4 | 4 |
| Feces | 2 | 0 | 1 |
| Perspiration | 1 | 0 | 0 |
| Fresh bread | 0 | 1 | 0 |
| Chemical smell | 0 | 0 | 1 |
| Metal | 0 | 0 | 1 |
| Coffee | 0 | 0 | 1 |
| None | 36 | 14 | 5 |
