## Supplementary material for "Olfactory, gustatory and trigeminal changes in non-hospitalized COVID-19 patients: an exploratory prospective cohort study": S2_Table

**S2 Table. Coping with smell and taste changes.**

**S2A** Coping with distorted smell

|  | **3 weeks**  (n=40) | **3 months**  (n=18) | **6 months**  (n=12) |
| --- | --- | --- | --- |
| No diminished smell | 33 | 12 | 2 |
| Nothing | 7 | 4 | 6 |
| Avoid bad smells | 0 | 1 | 4 |

**S2B** Coping with diminished smell

|  | **3 weeks**  (n=40) | **3 months**  (n=18) | **6 months**  (n=12) |
| --- | --- | --- | --- |
| No diminished smell | 2 | 0 | 4 |
| Nothing | 16 | 6 | 5 |
| Smell products to test if smell is back | 10 | 6 | 1 |
| Ask other people to smell rotten food | 6 | 2 | 1 |
| Pay extra attention to hygiene | 7 | 1 | 1 |
| Smell training | 3 | 5 | 2 |
| Ask help with cooking | 2 | 0 | 0 |
| Pay extra attention to expiration date | 2 | 0 | 0 |
| Clean more often | 1 | 0 | 0 |

**S2C** Coping with change in taste intensity

|  | **3 weeks**  (n=34) | **3 months**  (n=14) | **6 months**  (n=9) |
| --- | --- | --- | --- |
| Nothing | 22 | 6 | 4 |
| Experiment with stronger flavors | 6 | 4 | 1 |
| Ask help with cooking | 3 | 1 | 0 |
| Avoid certain flavors | 1 | 5 | 3 |
| Consume high moisturized foods | 1 | 0 | 0 |
| Taste products to test if taste is back | 1 | 0 | 0 |
| Chew longer | 1 | 1 | 0 |
| Mouth rinsing | 1 | 0 | 0 |
| Eating less | 0 | 1 | 1 |
| Sweets in mouth | 0 | 0 | 1 |
