## Supplementary material for "Olfactory, gustatory and trigeminal changes in non-hospitalized COVID-19 patients: an exploratory prospective cohort study": S3_Table

**S3 Table. Resemblance continuous taste**

|  | **3 weeks**  (n=9) | **3 months**  (n=5) | **6 months**  (n=3) |
| --- | --- | --- | --- |
| Something chemical | 0 | 2 | 0 |
| Metallic | 4 | 2 | 3 |
| Bitter | 1 | 1 | 0 |
| Sour | 1 | 0 | 0 |
| ‘Something odd’ | 1 | 1 | 0 |
| Infectious taste | 1 | 0 | 0 |
| Undescribable | 1 | 0 | 0 |
